# Large-scale genome- and phenome-wide analyses reveal the genetic architecture, pathophysiology and comorbidity of restless legs syndrome

**DOI:** 10.64898/2026.05.21.26353752

**Authors:** Xiaoxi Jing, Zhibin Wang, Jacob Williams, Junpeng Zhao, Kaiyan Ma, Xiaoxuan Fu, Jianxun Ren, Ouyang Chen, Hongen Kang, Jiecong Lin, Zeqi Yan, Jiaxiang Zhang, Shuo Zhang, Yafei Zhu, Hesheng Liu, Haoyu Zhang, Xihao Li, Yang Wu, Yi Tang, Yajie Zhao

**Affiliations:** Changping Laboratory, Beijing, China; Department of Neurology & Innovation Center for Neurological Disorders, Xuanwu Hospital, Capital Medical University, National Center for Neurological Disorders, Beijing, China; Division of Cancer Epidemiology and Genetics, National Cancer Institute, Rockville, MD, USA; Beijing Key Laboratory of Digital Medicine for Cognitive Disorders, Beijing, China; Academy for Advanced Interdisciplinary Studies, Peking University, Beijing, China; College of Future Technology, Peking University, Beijing, China; Department of Chemical Engineering, Tsinghua University, Beijing, China; Biomedical Pioneering Innovation Center, Peking University, Beijing, China; Department of Biostatistics, University of North Carolina at Chapel Hill, Chapel Hill, NC, USA; Department of Genetics, University of North Carolina at Chapel Hill, Chapel Hill, NC, USA; Institute of Rare Diseases, West China Hospital of Sichuan University, Chengdu, Sichuan, China

**Keywords:** Restless legs syndrome, Genome-wide association studies (GWAS), Phenome-Wide Association Study (PheWAS)

## Abstract

Restless legs syndrome (RLS) is a common sleep-related movement disorder, affecting approximately 4% of adults worldwide, with a substantially higher prevalence in individuals of European ancestry, reaching 5-13%. However, its genetic architecture and pathophysiology remain poorly understood. Here, we conducted a large-scale meta-analysis of more than 1.3 million individuals of European ancestry to explored the shared and ancestry-specific genetic architectures of RLS. We performed the first genome-wide association analyses of RLS in the UK Biobank and All of Us cohorts, leading to the identification of 15 novel RLS-associated loci. Using an updated variant-to-gene mapping and enrichment framework, we linked RLS risk signals to coherent neurodevelopmental and synaptic programs, with spatial transcriptomics analyses further localising these genetic effects to specific neural tissues. In addition, we performed the largest and most comprehensive phenome-wide association analysis of RLS to date, identifying over 100 diseases, lifestyle factors, and biomarkers significantly associated with RLS, thereby delineating its broad comorbidity spectrum. Finally, we conducted the largest brain MRI association analyses for RLS, providing further insight into how brain structure and connectivity are associated with the disorder. Together, these genome- and phenome-wide analyses clarify the genetic architecture, pathophysiology, and comorbidity landscape of restless legs syndrome.

## Introduction

Restless legs syndrome (RLS) is a common but frequently under-recognized sensorimotor disorder, affecting approximately 5-13% of individuals in Europe and North America and 1-3% in Asian populations^1^, with around 2-3% experiencing clinically significant symptoms requiring long-term pharmacological treatment, including dopaminergic agents, α2δ ligands or opioids^2^. RLS is characterized by an urge to move the legs, often accompanied by unpleasant sensory symptoms, that is triggered by rest or inactivity and transiently relieved by movement or external stimulation. Symptoms typically worsen in the evening or at night, leading to sleep disruption and substantial impairment in quality of life. RLS has been variably considered a comorbid manifestation or a potential prodromal feature of neurological, psychiatric and metabolic conditions, consistent with reported associations with neurodegenerative disease, psychiatric disorders, hypertension and cardiovascular disease^3^, as well as overlapping therapeutic responsiveness with movement disorders^2,4^. However, it remains unclear whether these associations reflect shared genetic architecture, downstream consequences of RLS, or residual confounding^5^.

Although RLS is widely regarded as a complex disorder shaped by genetic susceptibility, environmental exposures and their interactions, the biological mechanisms underlying disease risk and clinical heterogeneity remain incompletely understood^5^. Genetic studies provide an important entry point for elucidating these mechanisms. Twin and family studies indicate a substantial heritable component, with heritability estimates ranging from 40% to 70%^6,7^. Over the past decade, genome-wide association studies have identified an increasing number of common genetic variants associated with RLS, providing important insights into its genetic architecture^8-10^. In the recent large genome-wide association meta-analysis of RLS, common variants were estimated to explain approximately 20% of disease liability and 164 risk loci were identified, which advanced understandings about RLS genetics^10^. However, most existing studies have focused on common variation, leaving a substantial fraction of heritability unexplained and providing little insight into the cellular and molecular in which risk loci exert their effects.

Here, we presented a comprehensive genetic and phenotypic exploration of RLS by combining large-scale genome-wide association data with deep molecular, spatial and phenotypic analyses. Analysing more than 1.3 million individuals of European ancestry and approximately 0.2 million individuals of non-European ancestry across six studies, we refined the common-variant architecture of RLS and extended it through large-scale analysis of rare variants using genome sequencing data. By combining integrative variant-to-gene prioritisation with spatial transcriptomic mapping, we linked RLS risk signals to specific biological programmes and anatomical substrates. Leveraging the rich phenotypic resources of the UK Biobank, we investigated the clinical, behavioural and biological correlates of RLS. Using GWAS summary statistics from FinnGen, we assessed shared genetic architecture and causal relationships across thousands of traits. Finally, we systematically evaluated proposed links between RLS and neurodegenerative diseases. Together, this work provided an integrated framework for understanding the genetic architecture, pathophysiology and comorbidity of RLS.

## Results

We first leveraged recently released UK Biobank sleep questionnaire data, including the Cambridge-Hopkins Restless Legs Syndrome Questionnaire (CH-RLSq13), available for approximately 180,000 participants. Among 175,120 participants with valid questionnaire-derived RLS phenotypes, the mean age was 55.4 ± 7.6 years and 42.3% were male. We identified 13,662 individuals meeting criteria for RLS^11^, corresponding to a prevalence of 7.8%. Consistent with previous observations, RLS prevalence was approximately two-fold higher in women than in men (9.97% versus 4.84%). In UK Biobank, clinical coding captured only 1,916 RLS cases (ICD-10: G25.8), suggesting substantial under-ascertainment of RLS when relying on routine healthcare records. We therefore used the CH-RLSq13, a validated IRLSSG-based instrument, to define questionnaire-derived RLS for downstream analyses^9^ (Supplementary Table 1).

### Cross cohorts GWAS meta-analysis identifies 15 new risk signals for RLS

Using the newly derived questionnaire-derived RLS phenotypes, we firstly performed a genome-wide association study (GWAS) in unrelated individuals of European ancestry in the UK Biobank. We then combined these results with four additional cohorts in a genome-wide association meta-analysis of 1,130,977 individuals of European ancestry (41,268 cases and 1,089,709 controls) using METAL^12^, including EU-RLS-GENE, INTERVAL, FinnGen, and MVP (European ancestry only) (Fig. 1). In addition to the primary meta-analysis, we performed sensitivity meta-analyses excluding the UK Biobank when deriving polygenic risk scores (PRS) for UK Biobank participants, and excluding FinnGen when estimating genetic correlations within FinnGen, to avoid sample overlap and inflation of test statistics. Genetic correlations between cohorts were strong, with all pairwise estimates (*r_g_*) exceeding 0.88 (Supplementary Table 2, Fig. 2c), suggesting the consistency of the effect sizes across analysed cohorts. After quality control, 8,012,105 variants with minor allele frequency (MAF) ≥ 0.1% were included in the meta-analysis (Fig. 2a). We identified 9,487 genome-wide significant signals for RLS (*P* < 5x10^-8^). Independent association signals were defined using distance-based clumping combined with approximate conditional analysis, yielding 85 signals across 70 loci (Supplementary Table 3). To assess whether confounding factors such as population stratification exist, we applied LD score regression (LDSC). The SNP-based heritability estimated from the meta-analysis was 0.148 (0.015), and the LDSC regression intercept was 1.073 (0.011), suggesting that the observed genomic inflation was attributable to polygenic architecture rather than confounding.

**Fig. 1.**
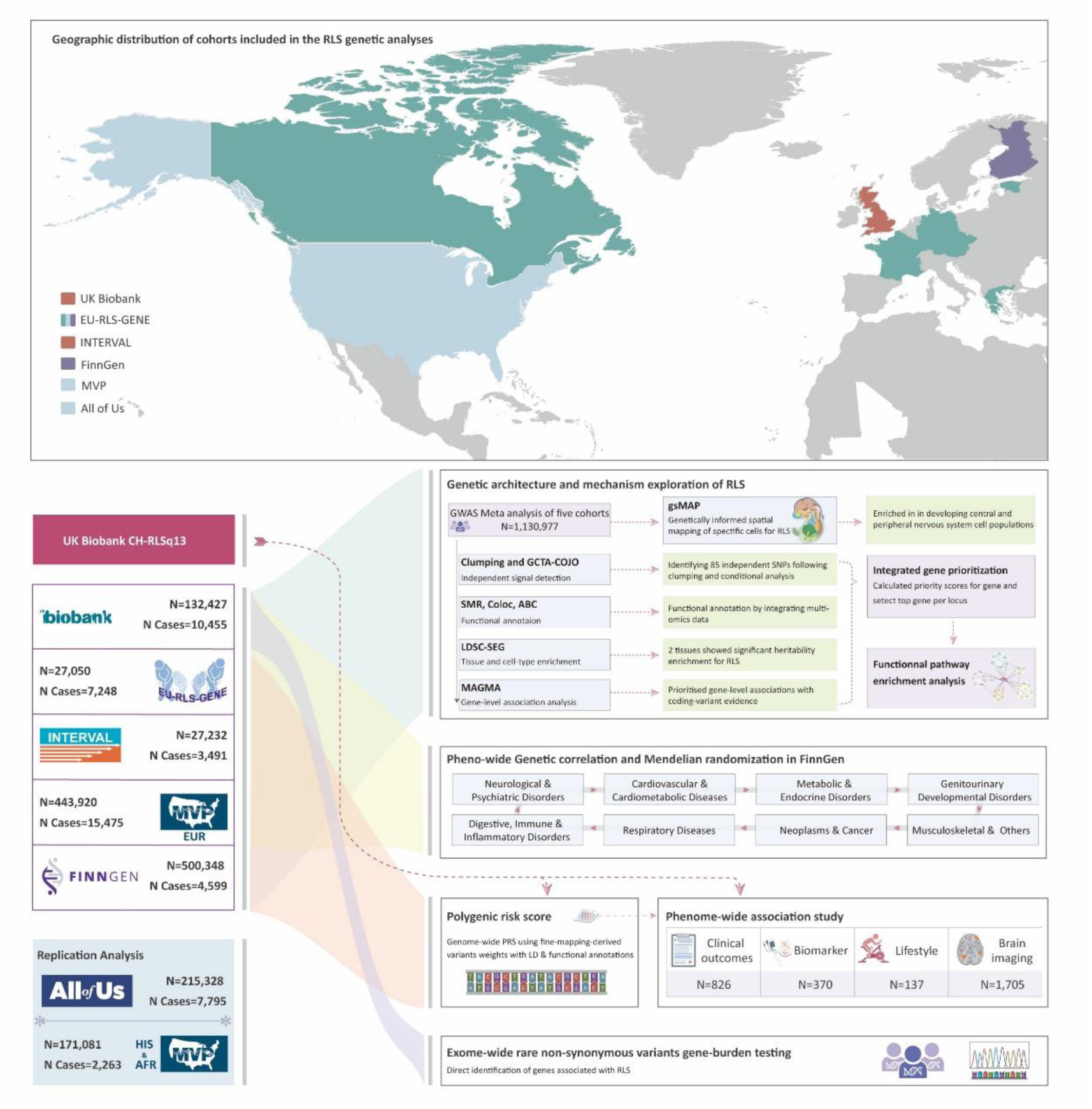
Global multi-cohort framework for elucidating the genetic architecture and biological mechanisms of restless legs syndrome (RLS). The upper panel illustrates the geographic distribution of cohorts included in the RLS genetic analyses, including UK Biobank, EU-RLS-GENE, INTERVAL, FinnGen, MVP, and All of Us. The lower panel outlines the integrative analytical pipeline used for genetic and multi-omics analyses.

**Fig. 2.**
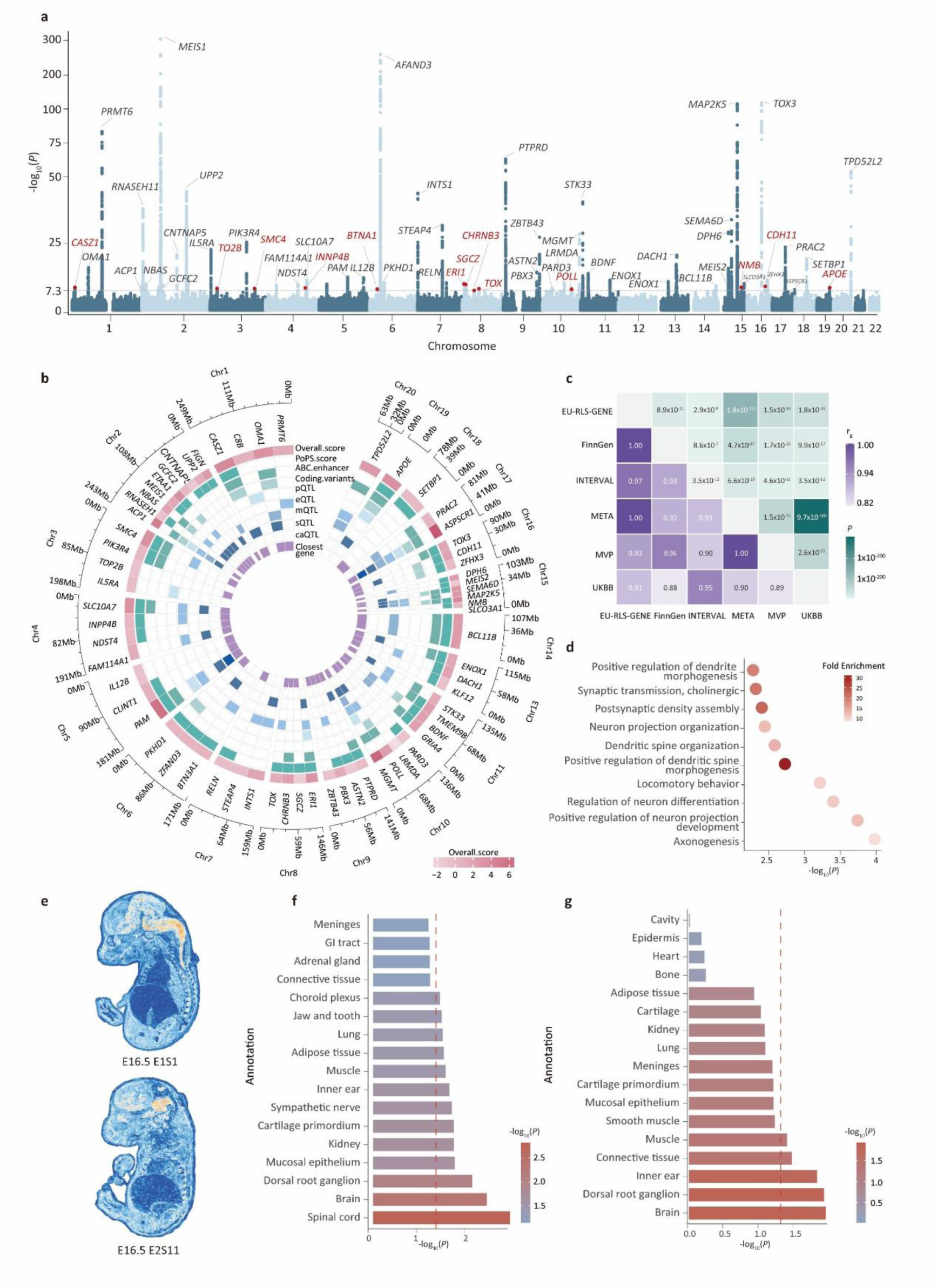
Multi-layered genetic and functional characterization of restless legs syndrome (RLS). a, Manhattan plot of the GWAS meta-analysis across five cohorts. Lead genes at each locus are annotated, with selected prioritized genes highlighted in red. b, Circular heatmap of GWAS-to-Genes (G2G) prioritization summary, which displays integrated evidence for candidate genes at each locus, including overall prioritization score, PoPS score, enhancer annotation, coding variants, pQTL, eQTL, mQTL, sQTL and caQTL. Color intensity reflects the relative strength of evidence for each annotation layer. c, Cross-cohort genetic correlation analysis among the five cohorts used for the GWAS meta-analysis. d, Gene Ontology (GO) pathway enrichment analysis based on G2G-prioritized genes. Color intensity indicates fold enrichment. e, Genetically informed spatial mapping (gsMap) of RLS onto embryonic mouse spatial transcriptomic atlases (E16.5: datasets E1S1 and E2S11). Heatmaps indicate spatial enrichment of RLS genetic risk across tissues. f-g, Tissue-level enrichment analysis corresponding to the two gsMap datasets shown in e.

We then compared our identifications with previously reported RLS loci. Of the 193 autosomal genome-wide significant loci reported by Schormair et al.^10^, 54 loci (27.98%) replicated at genome-wide significance (*P* < 5x10^-8^) in our meta-analysis. An additional 98 loci (50.78%) reached locus-level significance after Bonferroni correction (*P* < 0.05/193). In total, 159 loci (82.4%) showed nominal evidence of association (*P* < 0.05)^10^ (Supplementary Table 4). Extending the evaluation window to ±500 kb around the published lead variants increased the replication rate, with 189 loci (97.9%, *P*_Binom_ = 4.56x10^-53^) showing significant association signals (Supplementary Table 4). Based on publicly available summary statistics from the previous meta-analysis reported by Schormair et al. for the top 10,000 SNPs, 70 of our signals were located within ±500 kb of previously reported signals, whereas 15 represent novel associations (Supplementary Table 4)^10^.

To further validate our findings, we sought replication in the All of Us (AoU) cohort, which included 7,795 clinically diagnosed RLS cases and 207,533 controls. We then performed a GWAS in unrelated individuals of European ancestry in AoU. Of the 85 independent signals, 77 were available in AoU, of which 74 (96.1%) showed concordant directions of effect, and 46 reached nominal significance (*P* < 0.05) (Supplementary Table 5). To assess ancestry-specific genetic architecture, we extended replication analyses to non-European MVP cohorts (MVP-AFR and MVP-HIS). Concordance of effect directions was lower than in European cohorts, at 83.5% in MVP-HIS and 57.7% in MVP-AFR. Replication at nominal significance followed a similar pattern, with 32.9% of signals replicated in MVP-HIS and 12.9% in MVP-AFR (Supplementary Table 5). These findings indicate substantial ancestry-dependent heterogeneity, likely reflecting differences in allele frequencies, linkage disequilibrium structure, and effective sample sizes across populations.

To complement common-variant analyses, we conducted large-scale gene-based burden testing of rare coding variants for RLS using whole-genome sequencing data from 131,693 UK Biobank participants with both WGS data and questionnaire-derived RLS phenotypes. Gene burden tests were performed by collapsing rare variants (MAF < 0.1%) in each gene according to the following two overlapping predicted functional categories: (1) high-confidence protein truncating variants (HC PTVs) and (2) missense variants with AlphaMissense score > 0.564. However, no gene reached the predefined significance threshold (*P* < 2.2x10^-6^, Extended Data Fig. S1), indicating limited evidence for a contribution of rare coding variation to RLS risk at the current sample size.

**Fig. S1.**
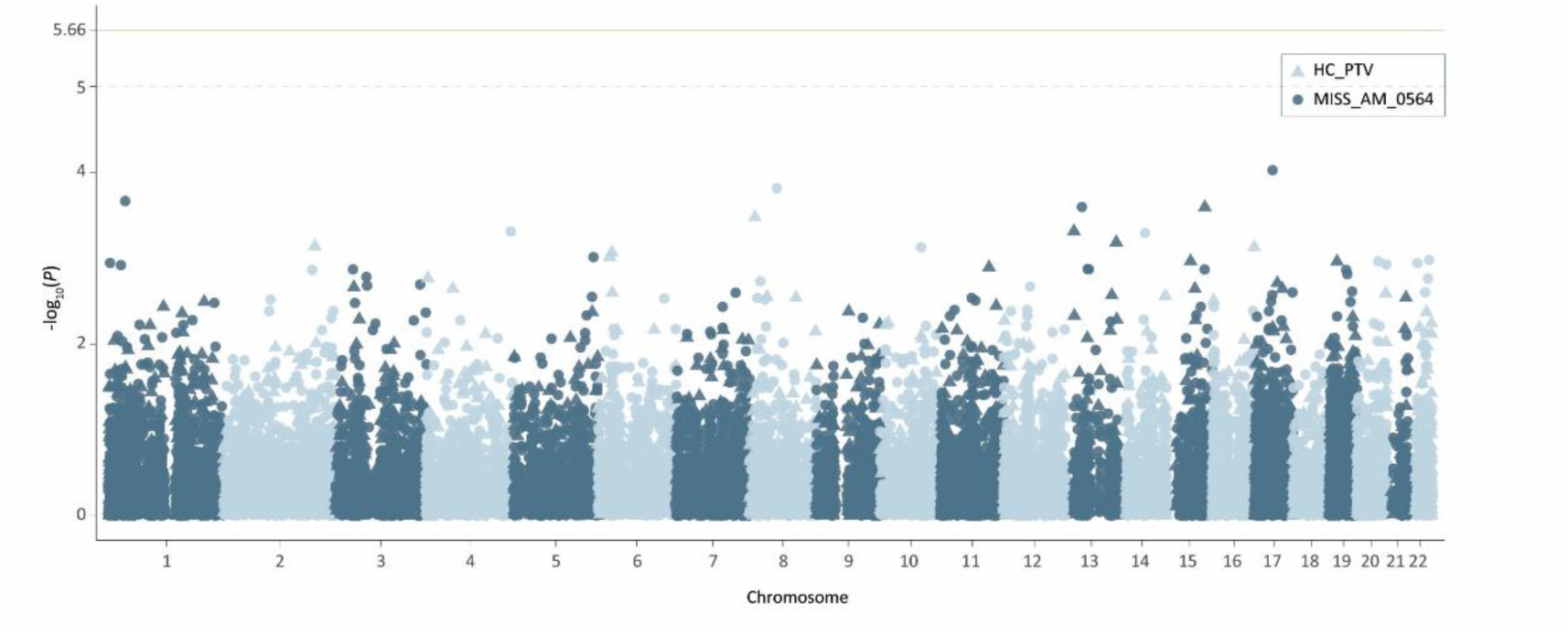
Manhattan plot of gene-based rare variant burden tests for restless legs syndrome (RLS). The red horizontal line indicates the exome-wide significance threshold, and the grey horizontal line denotes the suggestive significance threshold.

### Identifying RLS risk genes through variant-to-gene mapping

To prioritize putatively causal genes underlying the 85 RLS risk independent signals, we applied an updated multi-evidence GWAS-to-Genes (G2G) framework integrating genetic, regulatory, and functional information (Methods). Across signals, we leveraged multiple molecular quantitative trait datasets, including expression, protein, chromatin accessibility, splicing, and methylation QTLs, together with enhancer-gene links inferred using the Activity-by-Contact (ABC) model and coding variants in linkage disequilibrium with lead signals. In parallel, gene-level association analyses using MAGMA and PoPS provided complementary statistical support for gene prioritization. Mapping genes within ±500 kb of each lead variant yielded 486 candidate genes, which were evaluated using the integrated G2G evidence to identify genes most likely to mediate genetic risk for RLS.

Across all independent signals, we prioritised 64 high-confidence candidate genes for RLS, defined as the top-scoring gene per locus supported by at least three lines of evidence (Supplementary Table 6). Prioritised genes included established RLS candidates such as *MEIS1*, *PTPRD*, *SEMA6D*, and *GRIA4* (Supplementary Table 2, Fig. 2b). Among previously reported genes, *ASPSCR1* received strongest support across multiple molecular QTL classes, including brain cortex-specific sQTL and caQTL, and regulatory link evidence (Extended Data Fig. S2), consistent with a role in intracellular membrane-trafficking processes^13^. Newly prioritised genes included *CASZ1*, a zinc finger transcription factor implicated in neurodevelopmental programs^14^, and *CHRNB3*, encoding a nicotinic cholinergic receptor subunit^15^, highlightening potential contributions of transcriptional regulation and cholinergic signalling to RLS susceptibility. We also prioritized *APOE*, a key mediator of lipid transport and neurodegeneration^16^, raising the possibility that lipid-related pathways contribute to RLS biology. Notably, the signal at the *APOE* locus replicated across three large population cohorts (UK Biobank, FinnGen and MVP-EUR) but was not observed in smaller cohorts (EU-RLS-GENE and INTERVAL), which may reflect limited power and/or differences in phenotype definition or ascertainment.

**Fig. S2.**
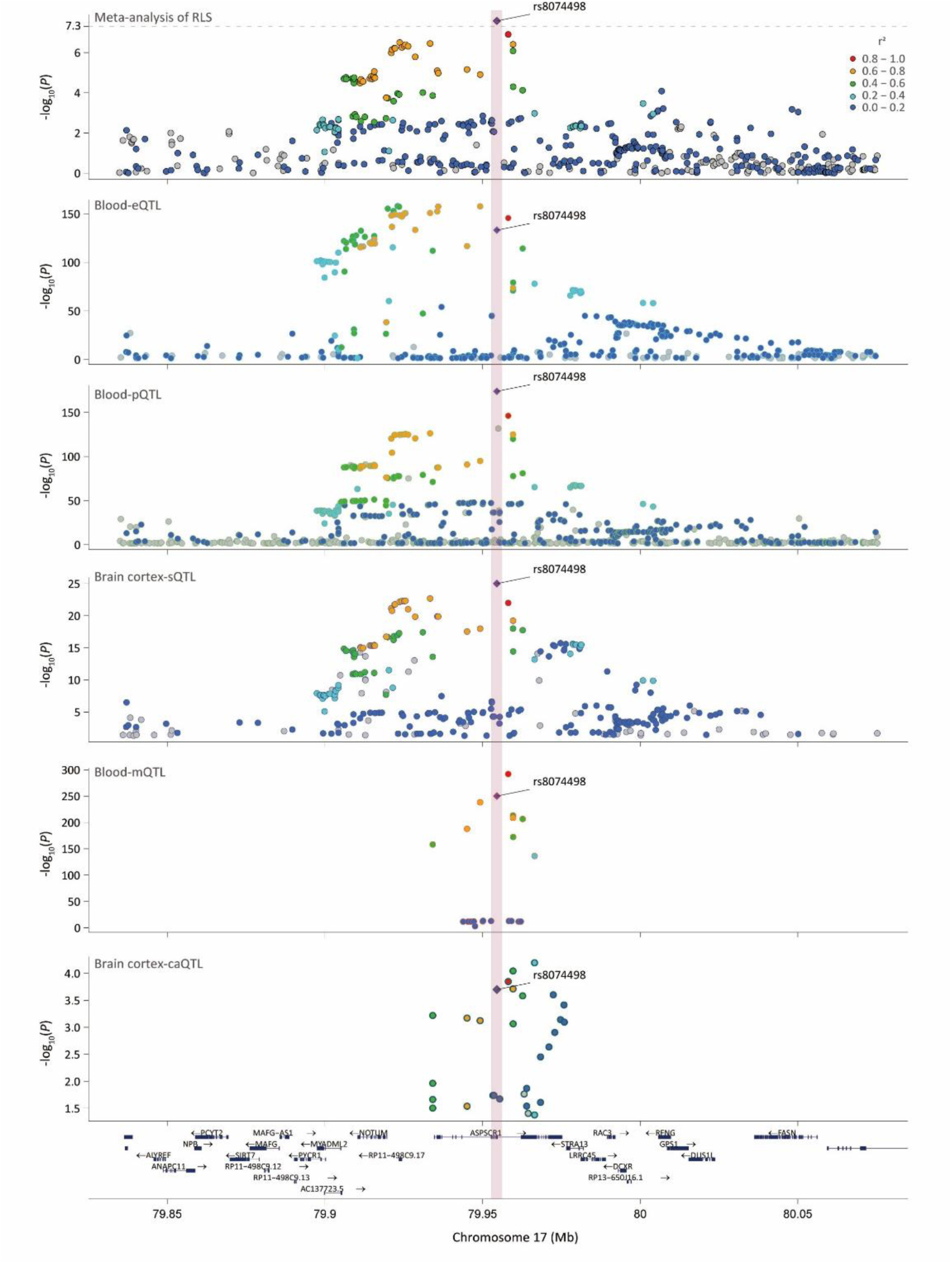
Regional association and molecular QTL signals at the rs8074498 locus. Regional association plots showing rs8074498 and corresponding molecular QTL associations in blood and brain cortex, including eQTL, pQTL, sQTL, mQTL, and caQTL.

### Biological pathways and functional enrichments/spatial and phenotypic convergence implicate sensory pathways in RLS

To investigate potential biological mechanisms, we performed enrichment analysis of G2G-prioritized genes using clusterProfiler. This analysis revealed consistent enrichment of biological processes related to neuronal development and structural organization. The most significantly enriched pathways included axonogenesis (*P* = 1.05x10^-4^), positive regulation of neuron projection development (*P* = 1.82x10^-4^), and regulation of neuron differentiation (*P* = 3.96x10^-4^), implicating mechanisms involved in the formation and maturation of neuronal projections (Fig. 2d, Supplementary Table 7). Additional enrichment was observed for pathways governing dendritic and synaptic architecture, including positive regulation of dendritic spine morphogenesis (*P* = 1.86x10^-3^), dendritic spine organization (*P* = 2.58x10^-3^), neuron projection organization (*P* = 3.54x10^-3^), and postsynaptic density assembly (*P* = 3.90X10^-3^), indicating a role in shaping neuronal connectivity and synaptic structure (Fig. 2d, Supplementary Table 7). Enrichment of cholinergic synaptic transmission (P = 4.73x10^-3^) further implicates specific neurotransmitter systems (Fig. 2d). Notably, enrichment of the behavioural process locomotory behaviour (*P* = 6.06x10^-4^) is consistent with abnormalities in sensorimotor circuits underlying the urge to move and the high prevalence of periodic limb movements in individuals with RLS^17^.

To explore the spatial cellular context of RLS risk, we projected meta-analysis signals onto a high-resolution spatial transcriptomic atlas from E16.5 mouse using the gsMap framework^18^. Across two independent sagittal datasets (E1S1 and E2S11), enrichment patterns were highly reproducible. The strongest signals localized to developing central and peripheral nervous system structures, including the spinal cord (Cauchy *P*_E1S1_=1.50x10^-3^), brain (Cauchy *P*_E1S1_=4.51x10^-3^, Cauchy *P*_E1S11_=1.16x10^-2^), and dorsal root ganglia (DRG, Cauchy *P*_E1S1_=9.05x10^-3^, Cauchy *P*_E1S11_=1.23x10^-2^), with additional enrichment in sympathetic nerve primordia (Cauchy P_E1S1_=2.36x10^-2^), cartilage primordium (Cauchy *P*_E1S1_=2.17x10^-2^), and muscles tissues (Cauchy *P*_E1S1_=3.02x10^-2^, Fig. 2f-g, Supplementary Table 8). Together, these spatial patterns place RLS genetic risk within a coherent developmental neuroanatomical framework, implicating early sensory-motor and autonomic circuits as primary substrates of disease susceptibility.

### Common variation influences the risk of phenotypic extremes

To evaluate the contribution of common genetic variants to RLS liability, we constructed a PRS in UK Biobank participants using a SBayesRC framework. We used SBayesRC because it incorporates functional annotations and enables genome-wide modelling of variant effects, thereby outperforming existing PRS methods. Posterior mean effect sizes were derived from the SBayesRC model trained on GWAS summary statistics from the meta-analysis excluding UK Biobank. After adjusting for age, sex, and the first ten principal components, the PRS explained an additional 3.27% of variance in RLS liability, as measured by Nagelkerke’s pseudo-R^2^. Participants were then ranked into 100 PRS percentiles, revealing a pronounced gradient of disease risk across the distribution. RLS prevalence increased from 1.12% in the lowest percentile to 21.83% in the highest percentile, corresponding to a 19.5-fold difference in risk between individuals at the extremes of polygenic burden (Fig. 3a). Predicted RLS prevalence closely matched observed prevalence. Individuals in the top 1% of the PRS exhibited a 2.91-fold increased risk of RLS (OR = 2.91, 95% CI: 2.69-3.14), whereas those in the bottom 1% showed markedly reduced risk (OR = 0.29, 95% CI: 0.25-0.35; Fig. 3b), highlighting the strong predictive power of PRS.

**Fig. 3.**
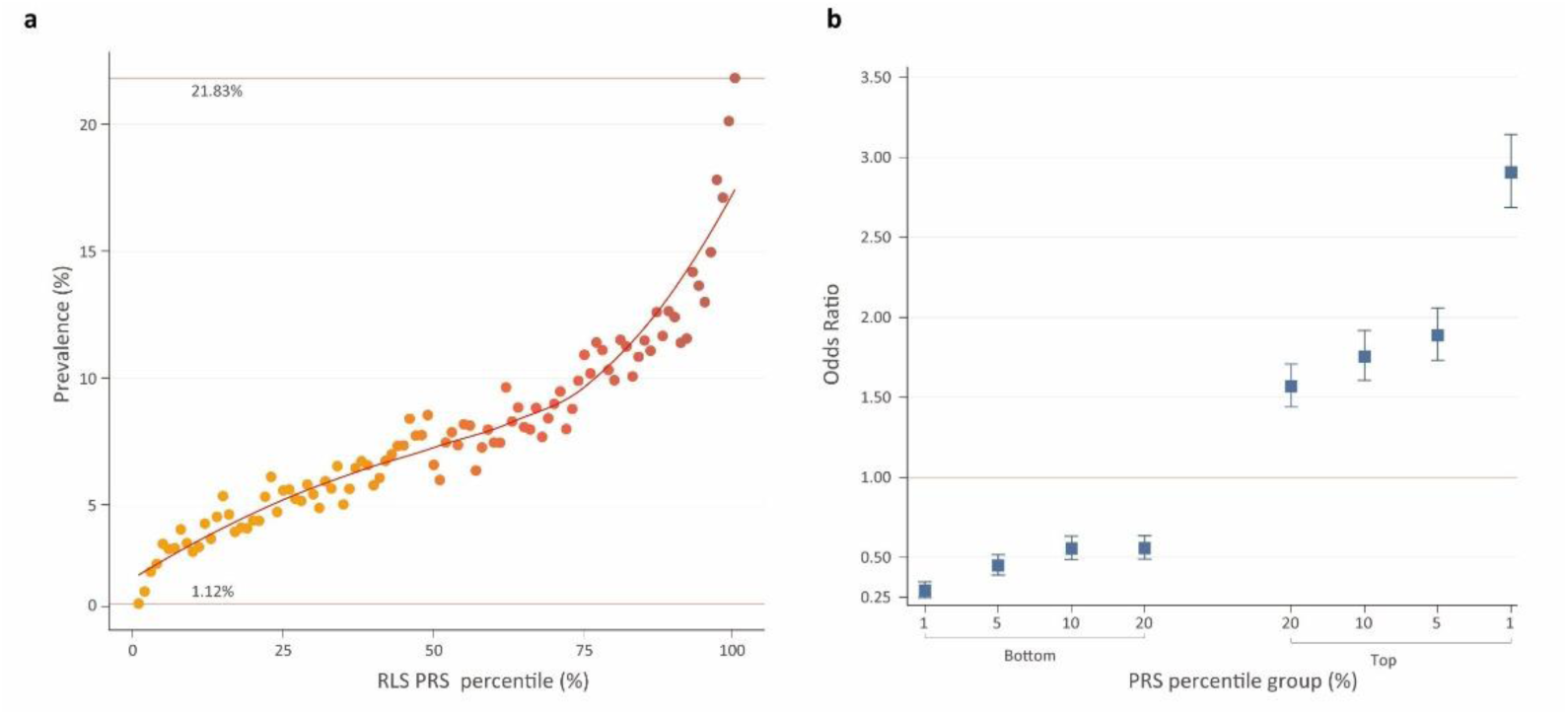
Polygenic prediction of restless legs syndrome (RLS) in the UK Biobank. a, Prevalence of RLS across polygenic risk score (PRS) percentiles in the UK Biobank. Individuals were ranked into 100 percentiles according to a genome-wide PRS for RLS. Each point represents the observed prevalence within a percentile. b, Estimated RLS risk in individuals in the top and bottom PRS percentile groups relative to the middle quintile (40-59%) of the PRS distribution. Odds ratios (ORs) were estimated using logistic regression adjusted for age, sex, and the first ten genetic principal components. The horizontal reference line denotes OR=1.

### Large scale phenome-wide analysis of RLS in UK Biobank

To investigate phenotypic associations of RLS, we conducted a systematic phenome-wide association study (PheWAS) in UK Biobank, encompassing a broad range of clinical outcomes, indicators and biomarkers, lifestyle factors, and measures of neuroimaging. Because fewer than 40% of participants had questionnaire-derived RLS phenotypes, we additionally performed a PheWAS for RLS PRS, allowing inclusion of a larger sample and enabling assessment of how genetic liability to RLS associates with other traits.

Clinical-outcome PheWAS was performed across 826 ICD-coded diseases, restricting analyses to conditions with at least 50 cases to ensure adequate statistical power. After Bonferroni correction, RLS was significantly associated with 49 clinical outcomes spanning a broad spectrum of disease domains. The strongest effect was observed for extrapyramidal and movement disorders (OR = 4.784, *P* = 4.41x10^-115^), consistent with concordance between questionnaire-derived and clinically ascertained RLS phenotypes. Other nervous-system disorders associated with RLS included migraine (OR = 1.297, *P* = 1.40x10^-12^) and nerve root and plexus compressions (OR = 1.501, *P* = 1.55x10^-8^) (Supplementary Table 9, Fig. 4a). RLS was also associated with multiple digestive-system disorders, including gastro-oesophageal reflux disease (OR = 1.408, *P* = 1.20x10^-36^) and irritable bowel syndrome (OR = 1.274, *P* = 6.15x10^-12^). Musculoskeletal and spinal conditions were likewise enriched, including other arthrosis (OR = 1.311, *P* = 1.36x10^-26^) and dorsalgia (OR = 1.259, *P* = 4.93x10^-17^) (Supplementary Table 9, Fig. 4a). Mental and behavioural disorders, including anxiety disorders (OR = 1.391, *P* = 5.55x10^-21^) and depressive episodes (OR = 1.294, *P* = 9.34x10^-18^), were significantly associated with RLS (Supplementary Table 9, Fig. 4a). Associations were also observed for asthma (OR = 1.260, *P* = 3.73x10^-17^) and cardiometabolic diseases, including primary hypertension (OR = 1.186, *P* = 5.80x10^-14^), chronic ischemic heart disease (OR = 1.299, *P* = 6.11x10^-10^), obesity (OR = 1.189, *P* = 1.65x10^-6^), and disorders of lipoprotein metabolism (OR = 1.118, *P* = 2.92x10^-5^) (Supplementary Table 9, Fig. 4a). Of the 48 clinical outcomes significantly associated with questionnaire-derived RLS, 38 were also significantly associated with the RLS PRS. Collectively, these findings highlight that RLS is linked to a distinct, multi-system disease landscape encompassing neurological, musculoskeletal, digestive, psychiatric, respiratory, and cardiometabolic domains.

**Fig. 4.**
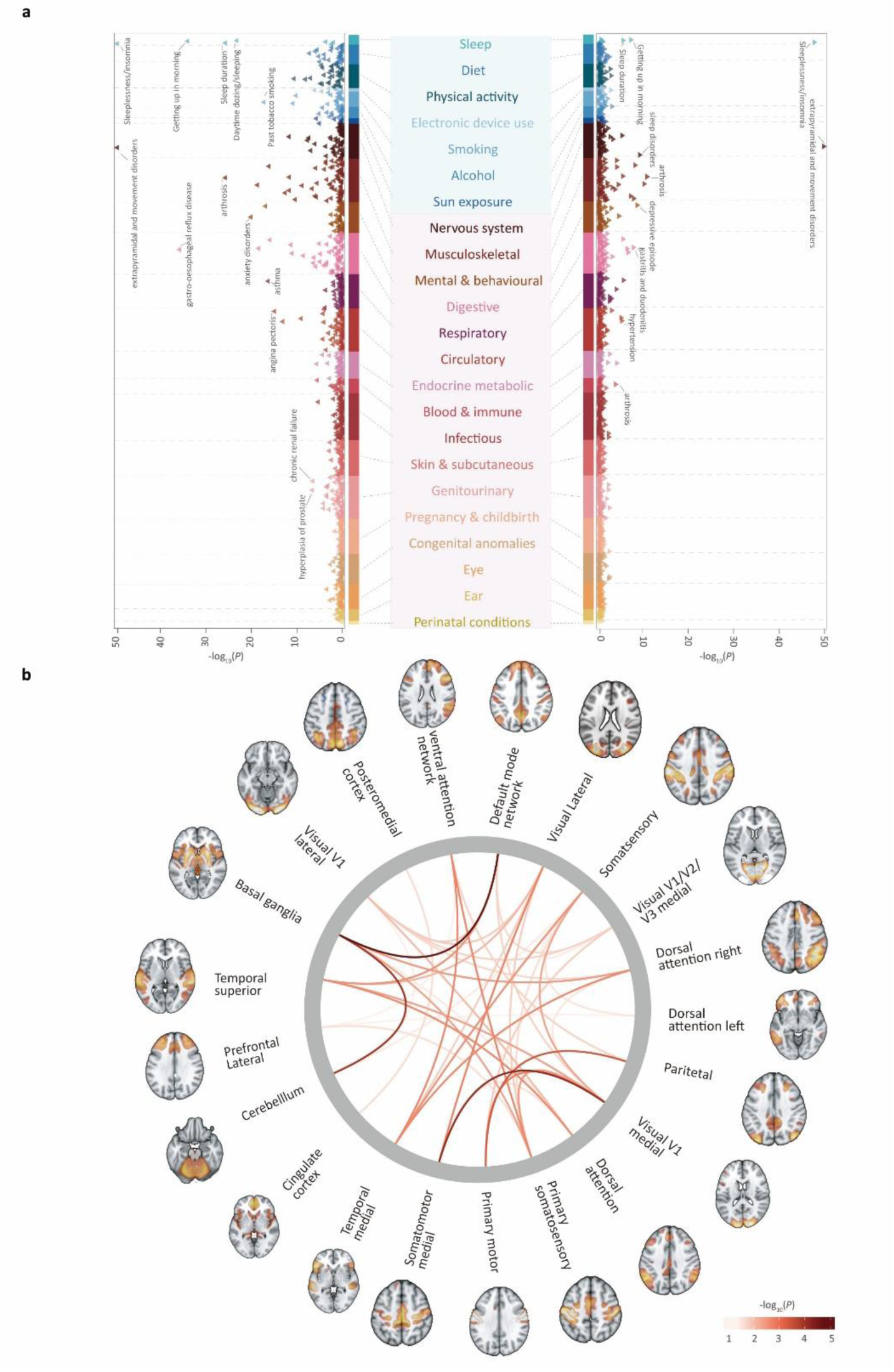
Phenome-wide associations of restless legs syndrome (RLS) and polygenic risk in UK Biobank. a, The left panel shows phenome-wide association results for questionnaire-derived RLS across a broad spectrum of clinical diagnoses and lifestyle-related traits. The right panel presents the corresponding PheWAS results for the RLS polygenic risk score (PRS). b, Brain phenome-wide association analysis linking RLS to functional connectivity patterns, derived from independent component analysis (ICA) of resting-state fMRI data and decomposed into 21 independent components.

**Fig. 5.**
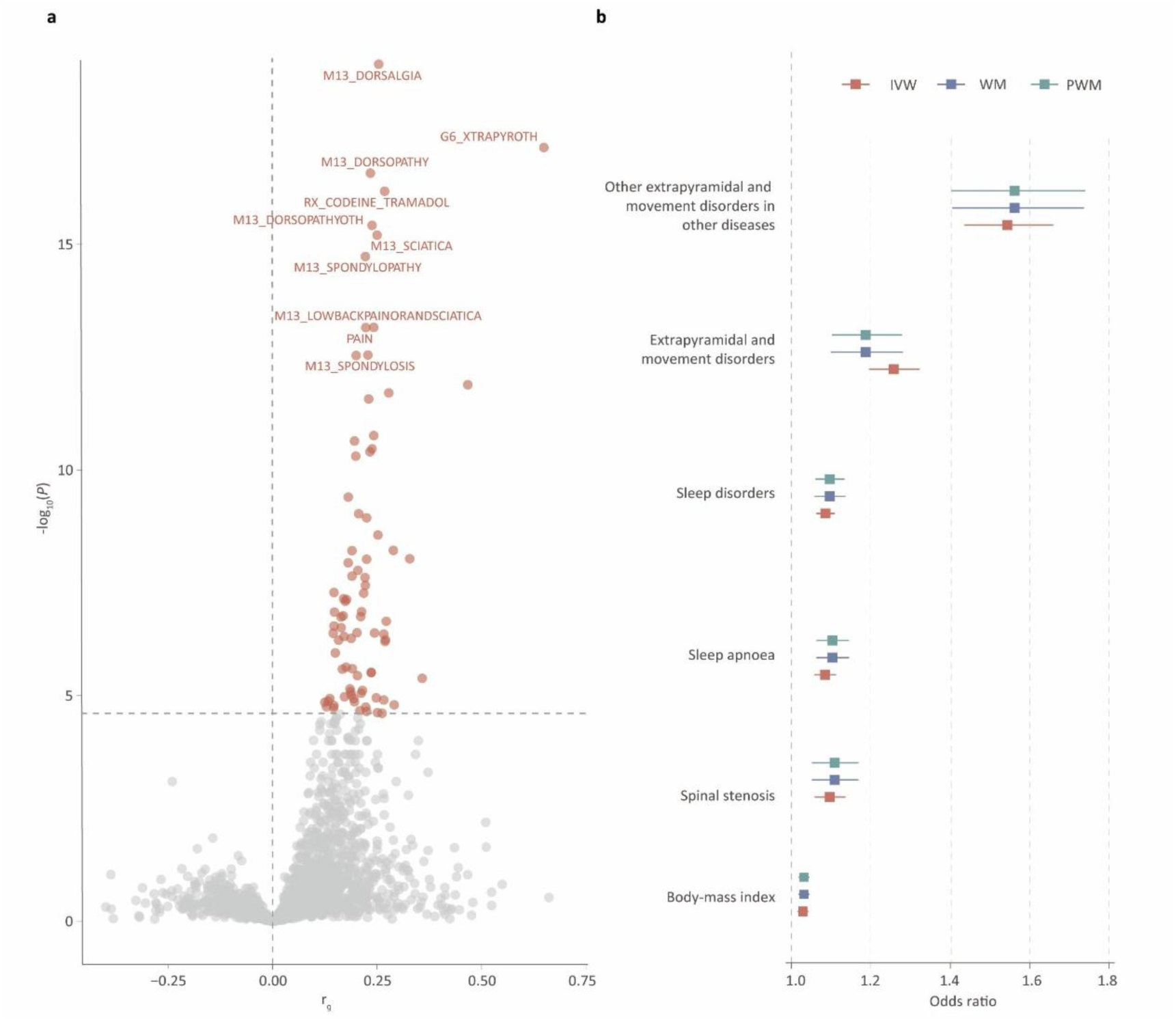
Genetic correlation and causal inference between restless legs syndrome (RLS) and FinnGen traits. a, Genome-wide genetic correlations between RLS and FinnGen clinical outcomes, estimated using linkage disequilibrium score regression (LDSC). b, Mendelian randomization (MR) analyses evaluating the potential causal effects of genetic liability to RLS on FinnGen outcomes. Odds ratios and 95% confidence intervals are shown for inverse-variance weighted (IVW), weighted median (WM), and penalized weighted median (PWM) models.

To characterise clinical indicators and biomarkers correlates of RLS, we tested associations between RLS status and 370 measures in the UK Biobank, encompassing body size measures, blood pressure, spirometry, blood biochemistry, urine assays, telomere length and NMR-based metabolomics (Supplementary Table 10). After multiple-testing correction, 16 indicators were significantly associated with RLS, 12 of which were also nominally associated with the RLS polygenic risk score. Higher RLS liability correlated with greater leg fat mass (OR_left_ = 1.047, *P*_left_ = 1.31x10^-5^, right: OR_right_ = 1.046, *P*_right_ = 2.31x10^-5^), elevated C-reactive protein (OR =1.049, *P* = 1.60x10^-5^), increased triglycerides in small VLDL particles (OR = 1.038, P = 5.27x10^-4^) and higher creatinine levels (OR = 1.039, *P* = 6.28x10^-4^), whereas total protein levels (OR = 0.951, *P* = 9.08x10^-6^) and resting pulse rate (OR = 0.959, *P* = 1.04x10^-4^) showed inverse correlations (Supplementary Table 10). We additionally observed a significant inverse association between circulating glucose levels (OR = 0.954, *P* = 4.32x10^-5^) and RLS status (Supplementary Table 10). However, this association was not supported by analyses using the RLS PRS, suggesting potential confounding or non-genetic influences. Consistent with previous reports, body mass index (OR = 1.034, *P* = 1.88x10^-3^) showed a nominally positive association with RLS (Supplementary Table 10).

### Lifestyle correlates of RLS

To characterise lifestyle correlates of RLS, we tested associations between RLS status and 137 lifestyle-related traits in the UK Biobank, encompassing sleep characteristics, smoking, alcohol consumption, diet, physical activity and related behaviours assessed at baseline. After Bonferroni correction, 30 traits were significantly associated with RLS. Sleep-related measures showed the strongest and most consistent associations, with five of seven sleep traits surpassing the significance threshold. RLS was positively associated with insomnia (OR = 1.539, *P* = 4.46x10^-185^), increased daytime dozing (OR = 1.237, *P* = 4.48x10^-24^) and chronotype (OR = 1.045, *P* = 1.50x10^-4^), and inversely associated with easier morning awakening (OR = 0.848, *P* = 7.11x10^-35^) and longer sleep duration (OR = 0.893, *P* = 1.35x10^-26^) (Supplementary Table 11, Fig. 4a). Beyond sleep, RLS was positively associated with smoking (OR = 1.116, *P* = 9.89x10^-12^), more frequent computer gaming (OR = 1.102, *P* = 5.56x10^-7^) and greater mobile phone use (OR = 1.039, *P* = 2.08x10^-5^), and inversely associated with higher fresh fruit intake (OR = 0.964, *P* = 8.50x10^-7^), use of sun or UV protection (OR = 0.946, *P* = 6.14x10^-6^), greater physical activity (OR = 0.996, P = 1.35x10^-11^) and lower sugar intake (OR = 0.869, *P* = 4.33x10^-6^) (Supplementary Table 11-12, Fig. 4a). Alcohol-related traits showed heterogeneous associations: greater average weekly consumption of champagne and white wine (OR = 0.987, *P* = 5.24x10^-6^) were inversely associated with RLS prevalence, whereas higher monthly spirits intake (OR = 1.062, *P* = 2.73x10^-4^) was positively associated (Supplementary Table 12, Fig. 4a). Together, these results indicate that RLS co-occurs with a wide range of lifestyle characteristics, particularly sleep-related behaviours.

### Neuroimaging insights into the pathophysiology of RLS

To investigate the neuroimaging correlates of RLS, we evaluated the associations between RLS and 1,705 brain imaging-derived phenotypes (IDPs) spanning structural imaging, functional imaging, susceptibility-weighted brain imaging and diffusion imaging, as reported by from Liu WS et al.^19^. After Bonferroni correction for multiple testing, 25 features were significantly associated with RLS. Subcortical motor and regulatory nuclei showed the strongest effects, led by multiple thalamic components, including the pulvinar (Pt, β_left_ = -0.086, P_left_ = 9.00x10^-9^, β_right_ = -0.075, P_right_ = 7.27x10^-8^), centromedian (CM, β_left_ = -2.788, P_left_ = 2.21x10^-7^, β_right_ = -2.471, P _right_ = 2.59x10^-6^), ventromedial (VM, β = -0.188, *P* = 1.05x10^-5^) and parafascicular (Pf) nuclei (β = -0.633, *P* = 1.38x10^-5^)^20,21^, all exhibiting significant volume reductions in RLS (Supplementary Table 13). Beyond subcortex, cortical associations were dominated by sensorimotor and higher-order association territories. Significant effects clustered in the posterior lateral fissure (β = -8.441 *P* = 1.40x10^-6^), inferior parietal lobule(particularly the supramarginal gyrus) (β = -46.350, *P* = 4.78x10^-7^), caudal middle frontal cortices (β = -27.044, *P* = 1.34x10^-5^), subcentral region (β = -11.728,*P* = 1.35x10^-5^) and the superior circular sulcus of the insula (β = -9.316, *P* = 1.70x10^-5^) (Supplementary Table 13). Next, we examined resting-state functional connectivity among 21 brain components derived from group independent component analysis (ICA) of large-scale networks exhibiting robust spatiotemporal coherence^22,23^. Strikingly, the resulting connectivity profile closely recapitulated the anatomical pattern observed in the IDP analyses: all significant edges linked core components of the thalamic–basal ganglia circuits with sensorimotor cortices and distributed association cortical networks. (Supplementary Table 13, Fig. 4b). After Bonferroni correction, significant alterations in RLS were observed in three network pairs. The default mode network-basal ganglia connection showed a negative association (β = -0.253, *P* = 2.16x10^-5^), consistent with the preceding structural findings. In contrast, the Visual V1 medial-Somatomotor medial (β = 0.289, *P* = 8.38x10^-5^) and Cerebellum-Basal Ganglia (β = 10.247, *P* = 1.39x10^-4^) connections exhibited positive associations, likely reflecting enhanced coupling within primary sensory-motor cortices and subcortical motor circuits in RLS. Together, these findings suggest that RLS involves structural and functional abnormalities in thalamo-basal ganglia circuits and in connected sensorimotor and association cortices.

### Genetic correlation and Mendelian randomisation across diseases and traits

To assess shared genetic architecture, we quantified genome-wide genetic correlations between RLS and 2,469 traits in FinnGen R12 using LDSC, based on GWAS summary statistics from the meta-analysis excluding FinnGen. After removing the invalid results and Bonferroni correction, 84 traits exhibited significant genetic correlations with RLS, broadly mirroring the phenotypic association patterns observed in UK Biobank. Significant correlations were observed for musculoskeletal and connective-tissue disorders, including dorsalgia (*r_g_* = 0.254, *P* = 1.02x10^-19^), low back pain with sciatica (*r_g_* = 0.242, *P* = 6.93x10^-14^), and spinal stenosis (*r_g_* = 0.196, *P* = 2.28x10^-11)^. Nervous-system disorders, including migraine (*r_g_* = 0.252, *P* = 2.76x10^-9^), digestive disorders such as gastro-oesophageal reflux disease(*r_g_* = 0.238, P = 3.40x10^-11^), psychiatric disorders anxiety (*r_g_* = 0.147, *P* = 5.18x10^-8^) and depression (r_g_ = 0.181, *P* = 3.99x10^-10^)), and respiratory conditions (asthma(*r_g_* = 0.187, *P* = 8.17x10^-6^), COPD (*r_g_* = 0.146, *P* = 1.87x10^-5^) also showed significant genetic overlap. Associations were observed for genitourinary disorders (other disorders of the urethra and urinary system (*r*_g_ = 0.218, *P* = 5.36x10^-8^), menorrhagia (*r_g_* = 0.213, *P* = 1.38x10^-7^)), visual disturbances (*r_g_* = 0.266, *P* = 1.25x10^-5^), and polycystic ovarian syndrome (*r_g_* = 0.194, *P* = 1.16x10^-5^), while cardiovascular traits showed nominal associations only. In UK Biobank, multiple sleep traits were significantly associated with RLS, and in FinnGen, significant positive genetic correlations were observed for insomnia (*r_g_* = 0.251, *P* = 2.38x10^-5^) and sleep apnoea (*r_g_* = 0.133, *P* = 1.32x10^-5^), indicating shared genetic liability between RLS and sleep disturbance. These results indicate that RLS shares broad, multi-system genetic architecture with conditions across neurological, musculoskeletal, digestive, psychiatric, respiratory, and genitourinary domains.

To determine whether shared genetic architecture translates into directional causal relationships, we performed a phenome-wide Mendelian Randomisation (MR) scan across the same FinnGen phenotypes. After correction for multiple testing, robust causal effects were observed primarily for sleep-related outcomes. Genetically predicted RLS was associated with an increased risk of composite sleep disorders (β_IVW_ = 0.082, *P* = 3.57x10^-8^) and sleep apnoea (β_IVW_ = 0.081-0.090; *P* = 2.93x10^-7^ to 6.15x10^-8^), with consistent directions of effect across all MR methods. No evidence of directional horizontal pleiotropy was detected based on MR-Egger intercept tests (*P*_intercept_ > 0.3). In contrast, reverse MR analyses using the IVW approach provided no evidence supporting causal effects of these traits on RLS liability (Supplementary Table 11). Consistent with previous reports implicating RLS in type 2 diabetes risk, we also observed a nominally significant association between genetically predicted RLS and type 2 diabetes (β_IVW_ = 0.059, *P* = 2.87x10^-4^).

### Evidence linking RLS to neurodegenerative diseases

Given the long-standing question of whether RLS predisposes individuals to neurodegenerative disease, we systematically assessed potential links with Alzheimer’s disease (AD) and Parkinson’s disease (PD) across multiple analytical layers. In UK Biobank PheWAS analyses, neither questionnaire-derived RLS nor the RLS polygenic risk score was significantly associated with clinically coded AD (*P* = 0.240-0.821, Supplementary Tables 9). Nominal associations were observed with PD (OR_RLS_ = 1.601, *P*_RLS_ = 0.009; β_RLS_PRS_ = 0.006, *P*_RLS_PRS_ = 0.02). However, analyses at the genetic level did not provide evidence of association or causal relationships. Linkage disequilibrium score regression based on the non-FinnGen meta-analysis revealed no significant genome-wide genetic correlation with AD (*P* = 0.205) or PD (*P* = 0.670), and Mendelian randomisation using FinnGen R12 summary statistics did not provide evidence for causal effects of RLS on AD (*P* = 0.791) or PD (*P* = 0.810), nor for AD or PD influencing RLS susceptibility (Supplementary Tables 15-16). Colocalization analyses between the 85 independent RLS signals and large AD and PD GWAS datasets identified a single high-confidence colocalised signal at the APOE region for AD (posterior probability for a shared causal variant, PP_4_ = 0.963, Supplementary Tables 17), while no convincing colocalization was observed for PD. These findings indicate that, despite nominal phenotypic associations, there is no strong genetic or causal evidence supporting a link between RLS and common neurodegenerative diseases.

## Discussion

Here, we conducted a large-scale European-focused GWAS of RLS, incorporating the first GWAS based on questionnaire-derived RLS phenotypes in the UK Biobank, four publicly available population-based GWAS datasets, and one RLS-specific study, two of which overlapped with previous RLS meta-analyses^10^. Although the total sample size was smaller than that of the most recent study, a substantial proportion of samples in that analysis were derived from 23andMe, which were not readily accessible. Our meta-analysis represented the largest publicly available GWAS resource for RLS, providing an important foundation for future genetic discovery and downstream functional studies in the field. We further performed the first GWAS of RLS in the All of Us cohort using newly released whole-genome sequencing data and observed robust replication of our findings. Across European-ancestry cohorts, we observed highly consistent genetic correlations despite differences in RLS case definitions, study designs, and environmental contexts, indicating a stable and shared common genetic architecture underlying RLS for European ancestry. In contrast, replication in non-European ancestry cohorts from MVP (AFR and HIS) was limited, likely reflecting differences in disease prevalence, allele frequency and linkage disequilibrium structure, as well as reduced effective sample sizes. Together, these findings highlight substantial ancestry-specific heterogeneity and underscore the need for larger, well-powered genetic studies of RLS in diverse populations.

By adopting a "variant-to-gene-to-pathway" framework^24^, we systematically integrated multi-omic cellular information, including expression, protein abundance, chromatin accessibility, splicing, and methylation quantitative trait loci, to elucidate the biological mechanisms underlying RLS. This integrative analysis revealed consistent enrichment of pathways related to neuronal development and structural organization. In addition, we performed first spatial mapping of RLS-associated variants, localising the strongest genetic signals to developing central and peripheral nervous system structures, including the spinal cord, brain, and dorsal root ganglia. These findings demonstrate the potential of integrating human GWAS results with spatial transcriptomic frameworks to pinpoint the most relevant anatomical regions and cell types underlying complex neurological phenotypes.

Although prior studies have examined associations between RLS and a broad range of clinical outcomes, lifestyle factors, biomarkers, and related traits in large population cohorts, these efforts have largely relied on PheWAS of individual risk variants, polygenic risk scores, or clinically underascertained RLS cases, limiting their ability to capture the full phenotypic spectrum of the disorder. By contrast, leveraging questionnaire-derived RLS phenotypes with improved case ascertainment in UK Biobank, integrating genome-wide polygenic risk scores with demonstrated predictive performance, and combining phenotypic association, genetic correlation, and Mendelian randomisation analyses across multiple large-scale cohorts, our study provides a more comprehensive and causality-aware delineation of the clinical and behavioural consequences of RLS genetic liability.

Our analyses further demonstrated both phenotypic and genetic associations between RLS and a broad range of diseases spanning neurological, musculoskeletal, digestive, psychiatric, respiratory, genitourinary, and cardiometabolic domains in the UK Biobank, enabled by the availability of enriched and comprehensive phenotypic data. A key limitation of these associations is the inability to infer temporal ordering, as RLS case status was derived from a recent online questionnaire, precluding determination of whether RLS onset preceded or followed the comorbid conditions. We also observed broad and pronounced effects of RLS on sleep quality, consistent with clinical observations that individuals with RLS frequently experience insomnia and involuntary periodic leg movements during sleep (PLMS) or wakeful rest (PLMW). Additionally, we found that well-established recommended lifestyle behaviours, such as higher fresh fruit intake and greater physical activity, were associated with a lower risk of RLS, whereas harmful behaviours, including smoking, were associated with an increased risk, highlighting potential modifiable risk factors for RLS. Although this study provides a comprehensive overview of associations between RLS and a wide range of clinical outcomes, lifestyle factors and biomarkers, RLS case status was derived from an online questionnaire administered more than 15 years after initial cohort enrolment. As a result, the analyses are susceptible to healthy participant bias, because individuals with severe morbidity or who had died were unable to complete the questionnaire. Consequently, some associations may be attenuated, and others should be interpreted with caution.

Neuroimaging provides a powerful approach to investigate the pathophysiology of RLS. However, previous studies comparing brain structure and connectivity between RLS cases and controls were limited to only a few hundred participants, restricting statistical power and increasing the chance of false-positive findings^25-29^. In contrast, leveraging the world’s largest whole-body imaging project conducted by UK Biobank, our study increased the sample size to over 35,000 participants, substantially enhancing the power to detect robust neuroanatomical and connectivity differences associated with RLS. Consistent with these earlier reports, but now at population scale, our analyses show that RLS is associated with coordinated alterations in both brain structure and functional connectivity, most prominently affecting thalamic-basal ganglia circuits^27,29^ and their sensorimotor and associative cortical projections^28,29^. These networks subserve sensory modulation, motor regulation, and arousal, offering a neurobiological framework for key clinical features of RLS, including abnormal sensory experiences, the urge to move, and sleep disruption. However, brain imaging was acquired years after cohort enrolment, and RLS status was determined only in a recent follow-up questionnaire. It therefore remains uncertain whether the observed brain differences reflect primary disease mechanisms or downstream consequences of long-standing symptoms and sleep disruption.

Prior studies have reported substantial clinical overlap between RLS and neurodegenerative disorders, with approximately 20% of individuals with PD^30^ and around 4% of those with AD exhibiting RLS^31^. These observations have raised long-standing concerns that RLS may predispose individuals to neurodegenerative disease. However, such estimates are derived largely from cross-sectional or clinic-based cohorts and are unable to establish temporal ordering, precluding determination of whether RLS precedes, follows, or merely co-occurs with neurodegeneration. To address this uncertainty, we examined the relationship between RLS and neurodegenerative disease using genetic approaches that are less susceptible to confounding and reverse causation. Across Mendelian randomisation and locus-level colocalization analyses, we found no evidence supporting a substantive genetic or causal link between RLS and either AD or PD. The single shared signal at the *APOE* locus is most parsimoniously interpreted as reflecting pleiotropic ageing-related effects rather than disease-specific biological overlap^32^. Together, these results suggest that the observed phenotypic associations between RLS and common neurodegenerative diseases are unlikely to reflect a strong shared genetic or causal basis.

Overall, this work advanced our understanding of genetic architecture, pathophysiology and comorbidity of RLS, which established a robust foundation for future mechanistic research, risk stratification, and the development of precision therapeutic strategies for RLS.

## Methods

### Ethics

The UK Biobank study has ethical approval from the North West Multi-centre Research Ethics Committee (REC reference: 13/NW/0157) as a Research Tissue Bank (RTB). This approval is renewed every five years, with successful renewals in 2016 and 2021. All participants provided informed consent, and the present analyses were conducted under UK Biobank application number 477126. The collection of participant information adhered to the AoU Research Program Operational Protocol (https://allofus.nih.gov/article/all-us-research-program-protocol). The AoU Institutional Review Board (IRB) (https://allofus.nih.gov/about/who-we-are/institutional-review-board-irb-of-all-of-us) is charged with reviewing the protocol, informed consent and other participant-facing materials for the AoU Research Program. For the remaining datasets, we used publicly available GWAS summary statistics for RLS obtained from the EU-RLS-GENE, INTERVAL, FinnGen, and Million Veteran Program (MVP) studies. All of these studies were approved by their respective institutional review boards or ethics committees.

### The definition of RLS in different cohorts

In UK Biobank, the RLS phenotype was defined using questionnaire items adapted from the validated Cambridge-Hopkins Restless Legs Questionnaire (CH-RLSq13; UK Biobank showcase IDs: 30579-30585, 32125, 32126). These items were included in the online follow-up survey, which contained over 160 questions related to sleep and was completed by approximately 180,000 participants. As of March 2025, this represents the largest sleep-questionnaire dataset available to biomedical researchers worldwide. RLS cases were identified as participants meeting all four core diagnostic criteria^11^: (i) an urge to move the legs, (ii) symptom onset or worsening during rest or inactivity, (iii) symptom relief with movement, and (iv) circadian variation. Individuals not meeting all RLS criteria were classified as controls, yielding 13,662 cases and 161,458 controls.

In the EU-RLS-GENE study, a multinational European case-control consortium established to investigate the genetics of RLS, 7,248 clinically diagnosed cases and 19,802 ancestry-matched controls were included. Participants were recruited through specialist neurology and sleep clinics across Europe, Canada, and the USA^10^. In the INTERVAL study, the Cambridge-Hopkins Restless Legs Questionnaire was used for phenotyping, and individuals with probable or definite RLS were combined to generate a binary RLS outcome, following the definitions established previously^8^, 3,491 RLS cases and 23,741 controls. In FinnGen, the RLS phenotype was based on clinical diagnosis (ICD-10 code G25.8) derived from primary care outpatient records, hospital discharge data, and cause-of-death registries (https://risteys.finngen.fi/endpoints/G6_RLS), resulting in 4,599 cases and 495,749 controls. In the Million Veteran Program (MVP), the RLS phenotype was similarly defined using clinically diagnosed cases based on ICD-9 code 333.94 and ICD-10 code G25.81 (https://phenomics.va.ornl.gov/web/cipher/phenotype-viewer/details?uqid=17f920c2b1624f04b55889e27eb4f1a9&name=Restless_Legs_Syndrome__Phecode_), yielding 15,475 cases and 428,445 controls.

### Genome-wide association testing of RLS in UK Biobank and meta-analysis

We applied the same GWAS framework as described in our previous analysis^8^ for UK Biobank. Genetic data were imputed centrally by the Wellcome Trust Centre for Human Genetics (WTCHG) using the combined Haplotype Reference Consortium (HRC) and 1000 Genomes reference panels, accessed via UK Biobank Data-Field 22828. In addition to UK Biobank’s standard central quality control, we restricted analyses to participants of white European genetic ancestry. This was determined using k-means clustering of the first four principal components derived from genome-wide variants genotypes. Individuals assigned to the white European cluster but who self-reported a non-European ancestry were excluded. To avoid confounding due to relatedness, only unrelated individuals were included in the association analyses. After all quality control filters, a total of 132,427 participants with both genotype and phenotype data were eligible for inclusion. Genome-wide association analysis was performed using REGENIE v3.3, implemented within the UK Biobank Research Analysis Platform and following recommended recommendations for UK Biobank analyses (https://rgcgithub.github.io/regenie/recommendations/). The association model included RLS status as the outcome variable, with genotyping chip, sex, age at questionnaire completion, and ten principal components of genetic ancestry included as covariates. GWAS summary statistics for RLS from the EU-RLS-GENE (N = 27,050) and INTERVAL (N = 27,232) studies were obtained from the GWAS Catalog (https://www.ebi.ac.uk/gwas/) under accession numbers GCST90399568-GCST90399573. Summary statistics for FinnGen (N= 500,348) and MVP RLS GWAS were obtained from FinnGen study (release R12) and MVP dbGaP repository phs002453 respectively. To maximise power for detecting association signals, we conducted a genome-wide meta-analysis across all RLS GWAS summary statistics described above using an inverse-variance weighted fixed-effects model implemented in METAL (release 2011-03-25). For the MVP dataset, only European-ancestry (N = 443,920) summary statistics were included. All datasets were harmonised for allele orientation and genomic build (GRCh37). In addition to the primary meta-analysis including all cohorts, we also performed two complementary analyses excluding either UK Biobank or FinnGen, respectively, for downstream analyses. To evaluate genomic inflation, we calculated the genomic inflation factor (λ_UK biobank_ = 1.103, λ_INTERVAL_ = 1.105, λ_EU-RLS-GENE_ =1.022, λ_FinnGen_ = 1.077, λ_MVP_ = 1.237), which indicated a slightly inflation for the meta-analysis. We estimated pairwise genetic correlations between cohorts using linkage disequilibrium score regression (LDSC, version 1.0.1). For LDSC analyses, we restricted variants to HapMap3 SNPs and used LD reference panels constructed from 1,000 Genomes Project European-ancestry samples.

### Genome-wide association testing of RLS in All of Us and replication analysis

In the All of Us (AoU) Research Program, RLS was defined based on clinically diagnosed cases identified using SNOMED code 32914008, ICD-9 code 333.94, and ICD-10 code G25.81 (https://databrowser.researchallofus.org/ehr/conditions/73754). Analyses were restricted to unrelated individuals of European genetic ancestry, as inferred by AoU, who either met the case definition for RLS or served as unaffected controls. After applying these criteria, the final dataset comprised 7,795 cases and 207,533 controls. Quality control (QC) procedures began with filters based on allele count-adjusted frequency (ACAF), the genetically inferred ancestry assignments provided by AoU, and the AoU-supplied list of samples with kinship coefficients > 0.1. To ensure adequate allele representation for downstream modelling, we applied an analysis-specific filter requiring minor allele count (MAC) > 30 prior to running REGENIE. REGENIE step 1 QC followed a hierarchical workflow. We first removed variants with missing call rate > 0.05 or minor allele frequency (MAF) < 0.05. Samples with missingness > 0.05 were then excluded to avoid unnecessary loss of variants. The remaining variant set was pruned for linkage disequilibrium using --indep-pairwise 1000 100 0.9 to generate an approximately independent marker set for ridge regression model fitting. REGENIE step 2 QC used thresholds tailored for association testing. Variants were filtered based on missing call rate > 0.05 and MAF < 0.1%, followed by removal of samples with missingness > 0.05. Association analyses were performed within the REGENIE framework using this QC’d dataset. To evaluate the concordance between the AoU findings and our meta-analysis results, we estimated pairwise genetic correlations using linkage disequilibrium score regression (LDSC, v1.0.1).

### Independent signals identification and gene prioritization for meta-analysis

To identify independent association signals and prioritise putative causal genes, we applied the updated G2G pipeline^24^ to variants included in the RLS GWAS meta-analysis summary statistics. Primary independent signals associated with RLS at genome-wide significance (*P* < 5x10^-8^) were first identified within non-overlapping 2-Mb windows, defined as ±1 Mb around the lead variant. Secondary independent signals were then detected using approximate conditional analysis implemented in GCTA v1.94.1^33^, with LD structures constructed from 25,000 randomly selected, unrelated UK Biobank participants of white European ancestry. Secondary signals were retained only if they (i) remained genome-wide significant after conditioning, (ii) exhibited low LD with the primary signal (r^2^ < 0.05), and (iii) showed minimal change in effect size upon conditioning. Variants with no protein-coding gene within 500 kb (based on NCBI RefSeq annotation) were excluded. In total, we identified 85 independent common susceptibility variants across 70 loci.

We mapped independent signals to putative causal genes using strategies consistent with those previously described^24^. Candidate genes were prioritised according to multiple evidence streams and scored based on cumulative support for causality. First, each lead signal was assigned to its nearest protein-coding gene. Second, signals and high-LD variants (r^2^ > 0.8) were annotated if they contained predicted deleterious coding mutations or if the corresponding gene demonstrated association through gene-level collapsing analyses of deleterious variants using MAGMA v1.1.0^34^. Third, non-coding signals and linked variants were annotated if they could be assigned to regulatory elements based on activity-by-contact enhancer maps^35^, restricted to cell and tissue contexts where the target gene was actively expressed. Fourth, we performed colocalization between GWAS signals and eQTLs using both the summary data-based Mendelian randomisation (SMR) with HEIDI testing (version 1.3.1)^36^ and an approximate Bayes factor implemented in R package ’coloc’ (v 5.2.3)^37^. These methods were applied jointly, as combined colocalization approaches have demonstrated improved robustness over single-method analyses^24^. Tissue-specific enrichment was evaluated using LD score regression applied to specifically expressed genes (LDSC-SEG)^38^, leveraging eQTL resources including Genotype-Tissue Expression (GTEx V7, available via https://gtexportal.org)^39^, eQTLGen^40^, and Brain-eMeta^41^. Equivalent analyses were performed for protein QTLs (pQTLs)^42^, chromatin-accessibility QTLs (caQTLs)^43^, splicing QTLs (sQTLs)^39^, and methylation QTLs (mQTLs)^44^. Finally, we integrated GWAS summary statistics with transcriptomic, pathway, and protein-protein interaction information using the polygenic priority score (PoPS) framework (v1.0)^45^ to nominate likely causal genes at each locus.

Once genes had been identified, we took forward the top ranked gene for each variant that had 2 or more lines of evidence to identify enriched pathways. We used R package ’clusterProfiler’, considering only those pathways and sets from the Gene Ontology project using the 61 prioritized genes identified through our G2G mapping framework. Biological process enrichment was assessed using the GO Biological Process (GO-BP) ontology, with significance evaluated using Fisher’s exact test.

### Cross-cohort effect consistency and ancestry comparison

To evaluate cross-cohort reproducibility of association signals, we assessed the consistency of effect directions and statistical significance for all 85 lead signals identified in the European meta-analysis (N = 1.13 million). For each of the five contributing European cohorts (UKB, EU-RLS-GENE, INTERVAL, FinnGen and MVP-EUR), we extracted per-cohort effect estimates and P value from GWAS summary statistics and compared them with the meta-analysis. Effect-direction concordance was calculated as the proportion of variants with the same sign of effect across datasets, and statistical significance replication was defined as the proportion of variants reaching nominal significance (*P* < 0.05) in each cohort. Binomial tests were performed to quantify the probability of observing the number of concordant variants under a null expectation of 50% agreement.

To examine ancestry-specific differences, we extended these analyses to additional ancestry groups within the MVP, including Hispanic/Latino (MVP-HIS) and African American (MVP-AFR) cohorts. Effect-direction concordance and nominal significance replication were computed as described above. Trans-ancestry genetic correlations (*r_g_*) between the European meta-analysis and each non-European cohort were estimated using LDSC, with standard errors and *P* values derived from the corresponding bivariate models. All comparisons were performed using harmonized variant sets matched by genomic position, effect allele, non-effect allele and allele frequency filters.

### Spatially resolved mapping of cells associated with RLS

We performed spatial transcriptomic enrichment analysis using gsMap version v1.73.5^18^, a framework that integrates gene-set signals with spatially resolved gene expression. RLS-associated genes were derived from our meta-analysis (N = 1,130,977), in which per-gene association statistics were aggregated using the GSS (gene set score) method. For spatial mapping, gene sets were projected onto the mouse embryo E16.5 whole-body spatial transcriptomic atlas generated by Stereo-seq (Sagittal sections E1S1 and E2S11). Significance was assessed using a permutation framework implemented in gsMap, with -log_10_(*P*) used to quantify regional enrichment. For visualization, we generated spatial score maps and tissue-level enrichment barplots based on curated embryonic anatomical annotations.

### RLS PRS derivation and validation in UK Biobank

We generated a RLS polygenic risk score from a meta-analysis of EU-RLS-GENE, INTERVAL, FinnGen, and MVP, comprising 41,268 cases and 1,130,977 controls. Variants-level posterior effect sizes for PRS construction were derived from genome-wide fine-mapping using the SBayesRC framework. The SBayesRC analysis was performed with GCTB^46^, which implements a Bayesian hierarchical mixture model that integrates genome-wide linkage disequilibrium (LD) patterns and functional genomic annotations to estimate the posterior effect size for each variant. Genome-wide LD was captured using precomputed eigen-decomposition reference panels provided with GCTB, ensuring consistency with the assumed genetic architecture. Functional annotations were incorporated from the comprehensive dataset published by Gazal et al.^46^, encompassing 96 genomic features relevant to complex trait architecture, including coding and conserved regions.

For PRS calculation, these SBayesRC-derived variants weights were applied to imputed genotype dosages of UK Biobank participants using PLINK2, generating centered and standardised per-chromosome scores that were summed to obtain a genome-wide PRS for each individual. In UK Biobank, PRS associations with RLS were first evaluated on a continuous scale. Associations were subsequently examined by PRS deciles, and by comparing individuals in the extreme PRS percentile groups (e.g., bottom 5% and top 5%) relative to the middle of the distribution, using logistic regression models adjusted for age, sex, and 10 genetic principal components.

### Phenome-Wide Association Study (PheWAS) of RLS in UK Biobank

To investigate phenotypic associations of RLS, we conducted a systematic PheWAS among UK Biobank participants who completed the sleep questionnaire. In addition to observed phenotypic associations, we evaluated the effects of the RLS polygenic risk score on other traits in all participants with genotype data. Continuous traits were analysed using linear regression, and binary traits using logistic regression adjusted for age, sex, and 10 genetic principal components, with Bonferroni correction applied for multiple testing within each trait group.

Clinical outcomes were defined using the "first occurrences" data in UK Biobank, which captures the earliest recorded occurrence of a given condition. These were derived from: (i) primary care data (Category 3000) mapped to 3-character ICD-10 codes from Read codes, (ii) hospital inpatient data (Category 2000) including ICD-9 and ICD-10 codes, (iii) death register records (Fields 40001 and 40002) with ICD-10 codes, and (iv) self-reported medical conditions (Field 20002) collected at baseline or follow-up visits. Diseases with fewer than 50 cases were excluded, leaving 826 clinically defined outcomes for PheWAS analyses.

To characterise clinical indicators and biomarker correlates of RLS, we systematically analysed 370 measures from UK Biobank Category 265, 100001 and 100006. These measures were collected during baseline visits at UK Biobank Assessment Centres using standardised protocols and encompass a broad spectrum of objective phenotypes including anthropometry, resting and exercise electrocardiography, blood pressure measurements, arterial stiffness indices, spirometry, hand-grip strength, bone densitometry of the heel, hearing tests, and carotid ultrasound metrics.

To assess the impact of lifestyle on RLS risk, we processed lifestyle-related traits from UK Biobank Category 100050. These include self-reported measures from the touchscreen questionnaire covering type and duration of physical activity, such as walking, moderate and vigorous exercise, and participation in strenuous sports.

We also analysed 1,705 brain imaging-derived phenotypes (IDPs) following the criteria established by Liu et al.^19^. Brain imaging processing, including artifact removal, cross-modality and cross-individual alignment, quality control, and phenotype extraction, follows the UK Biobank protocol (https://biobank.ndph.ox.ac.uk/showcase/showcase/docs/brain_mri.pdf). In addition to structural phenotypes, we examined 210 resting-state functional connectivity measures. The connectome was derived from 21 spatiotemporally coherent functional components obtained from an initial set of 25 independent component analysis (ICA) components, excluding four artefactual components^22,23^. Pairwise coupling strengths between these components were used as functional connectivity traits in the PheWAS.

### Genetic correlation and Mendelian Randomization (MR) between and RLS and 2,469 Traits in FinnGen

We leveraged GWAS summary statistics from the meta-analysis of UK Biobank, EU-RLS-GENE, INTERVAL, and MVP to systematically estimate the genetic correlation between RLS and 2,469 traits with available GWAS summary statistics in FinnGen R12 using LDSC v1.0.1. To investigate potential causal relationships, we performed bi-directional two-sample MR between RLS and these traits. Independent genetic instruments for each GWAS were selected using clumping and GCTA-COJO (GCTA v1.94.1), based on the UK Biobank European LD reference panel. Summary statistics were harmonized and analysed in R using the "TwoSampleMR" and "MendelianRandomization" packages. The inverse-variance weighted (IVW) estimator was used as the primary analysis, with weighted median, weighted mode, and MR-Egger regression applied as sensitivity analyses to evaluate robustness to horizontal pleiotropy. Heterogeneity and directional pleiotropy were assessed using Cochran’s Q statistic and the MR-Egger intercept, respectively, and MR-PRESSO was applied to detect and correct outlier variants. Results are reported as odds ratios per genetically predicted unit change in the exposure.

### Colocalization analysis between RLS and neurodegenerative disease

We performed colocalization analysis using the coloc package^37^ to identify genomic regions harbouring shared causal signals between RLS and neurodegenerative traits, including Alzheimer’s disease (AD) and Parkinson’s disease (PD). Colocalization was evaluated across loci that were significant in both RLS and the respective neurodegenerative trait. Evidence for a shared causal variant was defined as posterior probability of the H4 hypothesis > 0.75, indicating that both traits are associated and likely influenced by a single underlying causal variant within the region.

### Gene burden test for rare variants causing RLS

To identify rare germline variants (MAF < 0.1%) on coding genes associated with RLS risk, we performed gene-based burden tests across chromosomes 1-22 in 131,693 UK Biobank (UK Biobank) participants with both whole-genome sequencing and sleep questionnaire data. The WGS generation and processing pipeline for UK Biobank participants is described in previous study^47^. Briefly, 490,640 participants were sequenced at an average depth of 32.5x on the Illumina NovaSeq 6000. We annotated the variants with "FILTER = PASS" from ML-Corrected DRAGEN whole genome sequencing (WGS) in PLINK2 format using the ENSEMBL Variant Effect Predictor (VEP) v113.0, and analyses were restricted to rare non-synonymous variants (MAF < 0.1%). We defined protein-truncating variants (PTVs) as LOFTEE high-confidence stop-gained, splice donor/acceptor, or frameshift variants. Missense variants were further prioritised using AlphaMissense^48^, an AI-based pathogenicity prediction model, retaining putatively deleterious variants with AlphaMissense score > 0.564. Following established procedures^49,50^, we generated dummy genotype files in which each gene-mask combination was collapsed into a single pseudo-variant. Individuals carrying ≥1 qualifying variant in a gene were coded as heterozygous, irrespective of variant count. Dummy genotypes corresponding to rare HC-PTVs and deleterious missense variants (AM > 0.564) served as input for REGENIE v3.3 step 2, using identical model settings applied in the common-variant GWAS. Genes with < 30 qualifying variant carriers were excluded from each burden mask, yielding 8,451 genes for HC_PTV and 14,302 genes for MISS_AM_0564, totalling 22,753 tests. Accordingly, the exome-wide significance threshold was set using Bonferroni correction at: 0.05/22,753 = 2.2x10^−6^.

## Data availability

The UK Biobank phenotype and genome data described here are publicly available to registered researchers through the UK Biobank data access protocol. Information about registration for access to the data is available at https://www.ukbiobank.ac.uk/enable-your-research/apply-for-access. Summary statistics of the EU-RLS-GENE consortium GWAS and the INTERVAL GWAS are available at the GWAS Catalog (https://www.ebi.ac.uk/gwas/) under accession codes GCST90399568, GCST90399569, GCST90399570, GCST90399571, GCST90399572 and GCST90399573. GWAS summary statistics of FinnGen (Release 12) are available through the FinnGen consortium (https://r12.finngen.fi/) and statistics for MVP (EUR, HIS and AFR) are publicly available in dbGaP under accession number phs002453.

## Code availability

We provide information on publicly available software and settings in the Methods. Publicly available software used in this study includes PLINK v1.9 and PLINK v2.0, REGENIE v3.3, METAL (2011-03-25 release), GCTA v1.94.1, LDSC v1.0.1, GCTB, gsMap v1.73.5, MAGMA v1.1.0, SMR v1.3.1, PoPS v1.0, and the R statistical environment (v4.x), including the packages coloc v5.2.3, TwoSampleMR, MendelianRandomization, and clusterProfiler. Variant annotation was performed using Ensembl Variant Effect Predictor (VEP) v113.0 with the LOFTEE plugin. Rare-variant pathogenicity prediction was conducted using AlphaMissense.

## Acknowledgements

X.J., J.Z., K.M., X.F., J.R., H.K., Z.Y., S.Z., Y.Z., H.L., J.L., J.Zh. and Y.Z. are supported by Changping Laboratory (2024C-12-CJF). Z.W. was suppoted by National Natural Science Foundation of China (82401664). Y.W. was supported by the Fundamental Research Funds for the Central Universities, the 1·3·5 project for disciplines of excellence, West China Hospital, Sichuan University (ZYYC24006). Y.T. was supported by Noncommunicable Chronic Diseases-National Science and Technology Major Project (2025ZD0546200), National Natural Science Foundation of China (82220108009), Beijing Outstanding Young Scientist Program (JWZQ20240101023) and STI2030-Major Projects (2021ZD0201801).

This research was also supported in part by the Intramural Research Program of the NIH. The contributions of the NIH authors were made as part of their official duties as NIH federal employees, are in compliance with agency policy requirements, and are considered Works of the United States Government. However, the findings and conclusions presented in this paper are those of the authors and do not necessarily reflect the views of the NIH or the U.S. Department of Health and Human Services.

## Author contributions

All authors reviewed and contributed toward the drafting of the manuscript. X.J., Z.W., and J.Z. contributed toward the bioinformatics, genetic analyses and meta-analysis cross UK Biobank, EU-RLS-GENE, INTERVAL, FinnGen, and Million Veteran Program (MVP). J.W. contributed to genetic analyses of the All of Us data. K.M., X.F., J.R., O.C., H.K., J.L., Z.Y., J.Zh., S.Z., Y.Z., H.L. and H.Z. contributed to data analysis and/or interpretation. X.L., Y.W., Y.T. and Y.J.Z. designed and led the study.

